# Training needs of the aged care workforce providing services to Aboriginal and Torres Strait Islander peoples in South Australia: A cross-sectional survey

**DOI:** 10.64898/2025.12.21.25342785

**Authors:** Adriana Parrella, Jonathon Zagler, Anna Dawson, Tameeka Ieremia, Aidi Shuai, Graham Aitken, Natalie Warrior, Joni McArthur, Odette Pearson

## Abstract

**Objective(s):** The objective of this study was to explore the training needs and experiences of the aged care workforce providing services to Aboriginal and Torres Strait Islander peoples in South Australia.

**Method:** An online cross-sectional survey was distributed to aged care workers involved in the direct care or care coordination of Aboriginal and Torres Strait Islander peoples across South Australia between March and April 2024. Data collected included respondent demographics, aged care training experiences, needs and preferences. Descriptive and comparative analyses were conducted using R software.

**Results:** A total of 73 surveys were completed and analysed. Respondents reported a need for training across various topics, including social and emotional wellbeing (46.6%), grief and loss (45.2%), trauma-informed care (42.5%), and healthy aging (41.1%). Almost all respondents (98.6%) agreed or strongly agreed that training was important for skill development and delivering quality care. However, 39.7% reported not having received training specific to Aboriginal and Torres Strait Islander peoples in aged care. There was strong consensus on the importance of various training topics with most respondents rating them as *Extremely Important.* The two main barriers to participating in training were time constraints (58.9%) and the availability of training (46.6%). In-person training was the preferred delivery mode (45.2%) and case studies were the most preferred learning activity (78.1%).

**Conclusion(s):** Our data indicate there is an opportunity to strengthen cultural-specific training for aged care staff working with Aboriginal and Torres Strait Islander people in South Australia and highlight preferred training topics and delivery modes.

**Implications for Practice:** Findings have informed the development of *Walking Together in Aged Care,* a training program dedicated to building the capacity of the aged care workforce in meeting the needs of Aboriginal and Torres Strait Islander peoples. In particular, the findings guided training topics and content, delivery modes and interactive learning activities, and the access barriers that must be overcome.

**Implications for Policy:** Despite the *Royal Commission into Aged Care Quality and Safety* (2021) calling for improvements in the provision of workforce training, our results demonstrate a continued lack of training specific to supporting Aboriginal and Torres Strait Islander people in aged care.

## 1. Introduction

Aged care is Australia’s fastest growing sector, with an estimated 549,000 workers across residential, home and community settings.^1^ This workforce, comprising personal care workers, nurses, allied health professionals, administrative and ancillary staff, and informal carers, plays a critical role in delivering high-quality, safe care.^1^ The number of people employed within aged care is expected to increase further as Australia’s population continues to age, intensifying existing pressures relating to recruitment, retention, and capability.^2,3^

For Aboriginal and Torres Strait Islander peoples, the sovereign custodians and First Peoples of Australia, increasing life expectancy has led to increased need for and greater engagement with aged care services.^4^ However, access to health and aged care occurs within a context marked by colonisation, dispossession, and ongoing socio-political marginalisation, which have generated enduring inequities in health and ageing outcomes.^5^ As the population ages, chronic disease, comorbidity and multimorbidity presents at younger ages, often with aggressive disease progression leading to overall higher rates of these conditions than experienced by the non-Indigenous Australian population.^6^ The lived experiences prevalent and unique to the Aboriginal and Torres Strait Islander population and the ongoing intergenerational negative health and social impact of these, reinforce the need for culturally responsive and integrated aged care.

However, the Royal Commission into Aged Care Quality and Safety,^7^ in addition to a growing body of research,^8,9^ has highlighted that many Aboriginal and Torres Strait Islander peoples experience aged care as unsafe, disempowering and culturally alienating. This has since been reiterated by the Interim First Nations Aged Care Commissioner, who has emphasised the need for systemic reform, highlighting that many older Aboriginal and Torres Strait Islander peoples feel invisible within a system that was not designed for them.^10^

Such findings position workforce capability as a critical determinant of system reform. Within this context, the aged care workforce represents both a challenge and an opportunity for transformation. Aboriginal and Torres Strait Islander aged care workers are uniquely positioned to bridge this gap through their knowledge, experience and relationships with community.^10^ However, they remain underrepresented within the aged care system, accounting for only 1.2% of direct care staff across aged care programs in 2023.^1,11–13^ It is important to understand their training needs, learning preferences and barriers to develop training programs that will lead to an increase in Aboriginal and Torres Strait Islander aged care workforce who can deliver quality and safe care. Simultaneously, the non-Indigenous workforce often lacks the knowledge, skills and resources to understand and respond to the complex social, historical and cultural determinants of Aboriginal and Torres Strait Islander health and wellbeing.^14–18^ This dual challenge – of workforce underrepresentation and inadequate cultural capability – undermines the sector’s ability to deliver equitable aged care to Aboriginal and Torres Strait Islander peoples.

In response to the Royal Commission into Aged Care Quality and Safety, the strengthened Aged Care Quality Standards require aged care providers to demonstrate accountability for understanding and meeting the needs of Aboriginal and Torres Strait Islander peoples and explicitly acknowledge the need for a culturally safe workforce.^19^ However, evidence suggests that training opportunities remain inconsistent and fail to reflect the health, wellbeing and cultural needs of Aboriginal and Torres Strait Islander peoples,^14–18^ nor is there appropriate access to training that incorporates the needs of Aboriginal and Torres Strait Islander peoples.^15,16,20^

This study therefore seeks to extend the evidence on the training needs of the aged care workforce providing services to Aboriginal and Torres Strait Islander peoples. Sitting within a broader research project aimed at co-designing a training program that addresses the health, wellbeing and cultural needs of older Aboriginal and Torres Strait Islander peoples, This paper presents findings from a training needs assessment survey examining workforce training needs, delivery preferences, and participation barriers and enablers, with a specific focus on identifying differences in needs and preferences across workforce groups, including by ethnicity.

## 2. Methods

### 2.1 Study Design

This study was collaboratively designed by Wardliparingga Aboriginal Health Equity Theme (Wardliparingga) of the South Australian Health and Medical Research Institute and Aboriginal Community Services (ACS), an Aboriginal community-controlled aged care organisation and the largest provider of in-home and residential services to Aboriginal and Torres Strait Islander peoples within South Australia. This research builds on almost a decade of collaborative research activities. Together we developed a cross-sectional survey.

### 2.2 Ethics and Governance

The study received ethical approval by the South Australian Aboriginal Health Research Ethics Committee (#04-23-1084). Study activities were guided by a Project Steering Committee (PSC) which included Aboriginal community members receiving services from ACS, aged care workers employed by ACS, and members of the Wardliparingga research team. The PSC informed the research design, survey development, piloting, and interpretation and dissemination of findings. Throughout the research, we followed the nine principles for Aboriginal health research conduct in South Australia that were determined by Aboriginal and Torres Strait Islander communities.^21^

### 2.3 Survey Development

A cross-sectional survey was conducted with aged care workers involved in the direct care or care coordination of Aboriginal and Torres Strait Islander peoples. The survey, with closed and open-ended responses, was created on and distributed through REDCap (Research Electronic Data Capture). The survey was developed by the research team and the PSC. The items in the survey were designed by drawing upon existing questions, definitions, and categorisations used by the Australian Bureau of Statistics (e.g., age, gender, location, education), aged care workforce census reports ^11^, previous research undertaken by Wardliparingga,^14,15^ and the lived experiences of the PSC. The survey was piloted with the PSC and seven Wardliparingga staff, who were not on the research team, prior to distribution. The final survey included quantitative data collected using 52 closed-ended questions (i.e., 35 multiple-choice questions and 17 5-point Likert-scale items) and qualitative data collected from free-text comments available for 16 of the 52 questions. The survey collected information on 1) sociodemographic (i.e., age, gender, Indigenous status, employment role, education); 2) previous training; 3) training practices within respondents’ current workplace; 4) attitudes towards training, and 5) preferred training/skill development.

### 2.4 Participants

Staff who met all of the following inclusion criteria were invited to participate: 1) working in the direct care and/or coordination of aged care services for Aboriginal and Torres Strait Islander peoples in South Australia; 2) aged 18 years or over; 3) employed within non-government aged care organisations, independent for-profit aged care organisations, or Aboriginal community-controlled organisations (health and non-health); and 4) living in metropolitan, rural or remote South Australia.

### 2.5 Recruitment and Data Collection

A purposive sampling method was used for recruitment. First, a survey distribution list was compiled using information in GEN Aged Care, the South Australian Aboriginal Community Controlled Organisation Network, the National Aboriginal Community Controlled Health Organisation, and through sector contacts known to the research team and PSC. Next, potential participants were recruited via an email invitation to senior management of the selected organisations who were asked to distribute the study information and survey link to relevant employees. Potential participants were offered the opportunity to speak with the research team directly to ask questions or seek clarity over the research. To incentivise participation, respondents were offered an opportunity to enter a prize draw to win one of six $50 gift cards. The survey was conducted between 12 March and April 19, 2024. Participants were required to provide online informed consent prior to commencing the survey.

### 2.6 Data Analysis

Descriptive data analyses were performed in R (version 4.5.0). Postcode data were converted to Australian Statistical Geography Standard (ASGS) to classify respondents according to regional areas.^22^ Data were stratified by Indigenous status, as indicated by responses to “Question 4: Are you Aboriginal and/or Torres Strait Islander?”. For continuous variables such as hours worked, normality was assessed using the Shapiro-Wilk test, which indicated non-normal distribution. Consequently, the Kruskal-Wallis test was used for group comparisons. Ordinal variables, including age, education level, and attitudes towards training topics, were also analysed using the Kruskal-Wallis test. For categorical variables, Fisher’s Exact Test was applied when the minimum expected cell count was less than five. P-values were two-sided, with statistical significance set at p < 0.05.

## 3. Results

### 3.1 Sample

A total of 120 staff members attempted the survey, with 82 (68%) completions. Among the completed surveys, 9 respondents were excluded in the analysis, as they reported that their organisation did not provide aged care services to Aboriginal and/or Torres Strait Islander peoples. The remaining 73 respondents were included in the analysis.

Participant demographic characteristics are provided in Table 1. Most respondents were female (74.0%), born in Australia (74.0%), and based in major cities (67.1%) followed by very remote areas (19.2%). With regards to ethnicity, 60% of respondents identified as non-Indigenous, 26% identified as Indigenous and 14% preferred not to disclose their ethnicity. We report results by these three groups. A significant difference in geographic distribution was observed between the Indigenous and non-Indigenous workforce respondents (p < .001). Most non-Aboriginal respondents (86.4%) worked in major cities, while Aboriginal respondents were more widely dispersed, with the majority working in major cities (42.1%) and very remote areas (26.3%). The majority of total survey respondents were employed as coordinators or case managers (34.2%), followed by personal care workers or community support workers (23.3%). ‘Other’ roles included transport coordinator, liaison officer, and leisure and lifestyle coordinator.

**Table 1.** Participant demographics.

| Characteristics | All<br>N = 73<br>N (%) | Aboriginal<br>N = 19<br>N (%) | Non-Aboriginal<br>N = 44<br>N (%) | Prefer not<br>to disclose<br>N = 10<br>N (%) | p-<br>value* |
| --- | --- | --- | --- | --- | --- |
| **Gender** |  |  |  |  | .32 |
| Female | 54 (74.0) | 13 (68.4) | 34 (77.3) | 7 (70.0) |  |
| Male | 18 (24.7) | 6 (31.6) | 10 (22.7) | <5 (20.0) |  |
| Prefer not to say | <5 (1.4) | 0 (0.0) | 0 (0.0) | <5 (10.0) |  |
| **Age** |  |  |  |  | .15 |
| 18-24 years | <5 (5.5) | <5 (5.3) | <5 (4.5) | <5 (10.0) |  |
| 25-34 years | 24 (32.9) | 5 (26.3) | 15 (34.1) | <5 (40.0) |  |
| 35-44 years | 20 (27.4) | 5 (26.3) | 10 (22.7) | 5 (50.0) |  |
| 45-54 years | 14 (19.2) | <5 (15.8) | 11 (25.0) | 0 (0.0) |  |
| 55-64 years | 8 (11.0) | <5 (15.8) | 5 (11.4) | 0 (0.0) |  |
| 65 and over | <5 (4.1) | <5 (10.5) | <5 (2.3) | 0 (0.0) |  |
| **Country of birth:** |  |  |  |  | < .001 |
| Australia | 54 (74.0) | 19 (100.0) | 26 (59.1) | 9 (90.0) |  |
| Other | 19 (26.0) | 0 (0.0) | 18 (40.9) | <5 (10.0) |  |
| **Geographic region** |  |  |  |  | < .001 |
| Major Cities of Australia | 49 (67.1) | 8 (42.1) | 38 (86.4) | <5 (30.0) |  |
| Outer Regional Australia | 5 (6.8) | <5 (10.5) | <5 (6.8) | 0 (0.0) |  |
| Remote Australia | 5 (6.8) | <5 (21.1) | 0 (0.0) | <5 (10.0) |  |
| Very Remote Australia | 14 (19.2) | 5 (26.3) | <5 (6.8) | 6 (60.0) |  |
| **Occupation:** |  |  |  |  | .16 |
| Personal Care / Community Support Worker | 17 (23.3) | 6 (31.6) | 10 (22.7) | <5 (10.0) |  |
| Coordinator/Case Manager | 25 (34.2) | 8 (42.1) | 11 (25.0) | 6 (60.0) |  |
| Service/Program Manager | <5 (5.5) | 0 (0.0) | <5 (9.1) | 0 (0.0) |  |
| Allied Health Professional | <5 (4.1) | 0 (0.0) | <5 (4.5) | <5 (10.0) |  |
| Nurse | 7 (9.6) | 0 (0.0) | 6 (13.6) | <5 (10.0) |  |
| Aboriginal Health Practitioner | <5 (2.7) | <5 (10.5) | 0 (0.0) | 0 (0.0) |  |
| Administration | 5 (6.8) | 0 (0.0) | 5 (11.4) | 0 (0.0) |  |
| Ancillary roles | <5 (1.4) | 0 (0.0) | <5 (2.3) | 0 (0.0) |  |
| Medical Professional | <5 (1.4) | <5 (5.3) | 0 (0.0) | 0 (0.0) |  |
| Others | 8 (11.0) | <5 (10.5) | 5 (11.4) | <5 (10.0) |  |
| **Highest education completed:** |  |  |  |  | .44 |
| Secondary education - year 9 and below | <5 (1.4) | <5 (5.3) | 0 (0.0) | 0 (0.0) |  |
| Secondary education - year 10 and above | 21 (28.8) | 8 (42.1) | 9 (20.5) | <5 (40.0) |  |
| Certificate III or IV | 15 (20.5) | 5 (26.3) | 8 (18.2) | <5 (20.0) |  |
| Diploma or Advanced Degree | 14 (19.2) | <5 (10.5) | 9 (20.5) | <5 (30.0) |  |
| Bachelor's Degree | 14 (19.2) | <5 (15.8) | 10 (22.7) | <5 (10.0) |  |
| Graduate Certificate or Diploma | <5 (2.7) | 0 (0.0) | <5 (4.5) | 0 (0.0) |  |
| Masters/PhD | 6 (8.2) | 0 (0.0) | 6 (13.6) | 0 (0.0) |  |
| **Qualification completed in Australia (n=53) †** | 52 (98.1) | 10 (100.0) | 36 (97.3) | 6 (100.0) | > .99 |
| **Type of organisation** |  |  |  |  | .002 |
| Aboriginal community-controlled (non-health) | 25 (34.2) | 9 (47.9) | 8 (18.2) | <5 (10.0) |  |
| Aboriginal community-controlled health organisation | 19 (26.0) | 7 (36.8) | 11 (25.0) | 7 (70.0) |  |
| Non-government-organisation/s | 24 (32.9) | <5 (15.8) | 20 (45.5) | <5 (20.0) |  |
| Independent/for-profit | 5 (6.8) | 0 (0.0) | 5 (11.4) | 0 (0.0) |  |
| **Type of care** |  |  |  |  | .48 |
| Residential care | 12 (16.4) | <5 (10.5) | 9 (20.5) | <5 (10.0) |  |
| In-home care | 42 (57.5) | 10 (52.6) | 25 (56.8) | 7 (70.0) |  |
| Both residential and in-home care | 13 (17.8) | <5 (15.8) | 8 (18.2) | <5 (20.0) |  |
| Other | 6 (8.2) | <5 (21.1) | <5 (4.5) | 0 (0.0) |  |
| **Among those who provide in-home care (n = 55), type of in-home program ‡** |  |  |  |  | .04 |
| Commonwealth Home Support Programme (CHSP) | 9 (16.4) | <5 (30.8) | <5 (12.1) | <5 (11.1) |  |
| Home Care Package Program (HCP) | 21 (38.2) | 6 (46.2) | 9 (27.3) | 6 (66.7) |  |
| Both CHSP and HCP | 25 (45.5) | <5 (23.1) | 20 (60.6) | <5 (22.2) |  |
| **Employment** |  |  |  |  | .04 |
| Casual | 16 (21.9) | <5 (10.5) | 8 (18.2) | 6 (60.0) |  |
| Permanent | 51 (69.9) | 15 (78.9) | 32 (72.7) | <5 (40.0) |  |
| Fixed term | <5 (4.1) | <5 (10.5) | <5 (2.3) | 0 (0.0) |  |
| Independent subcontractor | <5 (4.1) | 0 (0.0) | <5 (6.8) | 0 (0.0) |  |
| **Years in current role** |  |  |  |  | .90 |
| Less than 1 year | 22 (30.1) | 7 (36.8) | 14 (31.8) | <5 (10.0) |  |
| 1-2 years | 24 (32.9) | 6 (31.6) | 12 (27.3) | 6 (60.0) |  |
| 3-4 years | 14 (19.2) | <5 (10.5) | 9 (20.5) | <5 (30.0) |  |
| 5-10 years | 6 (8.2) | <5 (5.3) | 5 (11.4) | 0 (0.0) |  |
| More than 10 years | 7 (9.6) | <5 (15.8) | <5 (9.1) | 0 (0.0) |  |
| **Years working with Aboriginal and Torres Strait Islander aged care clients** |  |  |  |  | .51 |
| Less than 1 year | 16 (21.9) | <5 (10.5) | 14 (31.8) | 0 (0.0) |  |
| 1-2 years | 20 (27.4) | 7 (36.8) | 7 (15.9) | 6 (60.0) |  |
| 3-4 years | 16 (21.9) | <5 (15.8) | 9 (20.5) | <5 (40.0) |  |
| 5-10 years | 10 (13.7) | <5 (10.5) | 8 (18.2) | 0 (0.0) |  |
| More than 10 years | 11 (15.1) | 5 (26.3) | 6 (13.6) | 0 (0.0) |  |
| **Number of Aboriginal and/or Torres Strait Islander aged care clients, Median (IQR) §** | 20 (7, 50) | 25 (12, 60) | 20 (3, 55) | 15 (9, 15) | .32 |
| **Work hours per week, Median (IQR) §** | 38 (33, 38) | 38 (35, 38) | 38 (30, 39) | 36 (30, 38) | .57 |

Data were presented as n (%) unless stated otherwise. \* Medians and percentages were compared using Kruskal-Wallis test and Fisher's Exact test, respectively. Ordinal variables, including age, highest education completed, years in current role, and years working with clients were analysed using the Kruskal-Wallis test. † 53 respondents with higher education than Secondary education. ‡ 55 respondents support clients in in-home care. § IQR 25th–75th percentile.

There was a significant difference in the type of organisation participants worked in by Indigenous status (p = .002). Most Aboriginal respondents (47.9%) worked in Aboriginal community-controlled (non-health) organisations, most non-Aboriginal respondents (45.5%) worked in non-government organisations, and most respondents (70.0%) who preferred not to disclose their Indigenous status worked in Aboriginal community-controlled health organisations.

The majority of respondents were employed within in-home care (57.5%) followed by a combination of residential and in-home care roles (17.8%) (p = 0.04). Most Aboriginal respondents (78.9%) and most non-Aboriginal respondents (72.7%) were employed on a permanent basis, while most respondents who preferred not to disclose their Indigenous status (60.0%) were employed on a casual basis (p = 0.04).

### 3.2 Previous Training Experience and Training Needs

Table 2 outlines responses regarding training experience and training needs. Most respondents indicated they were required to complete training upon starting their role at their organisation (78.1%) and were paid to participate in training (89.0%). Of the 16 respondents who were not required to undergo training, 56.3% believed they should have received it. Additionally, 32.9% of respondents reported annual training requirements while 34.2% were unsure about the required training frequency within their workplace. Specific training received by respondents included workplace policies and procedures, cultural awareness, and manual handling.

**Table 2.** Previous training experience and training needs (n=73)

|  | Overall<br>N = 73<br>N (%) | Aboriginal<br>N = 19<br>N (%) | Non-Aboriginal<br>N = 44<br>N (%) | Prefer not<br>to disclose<br>N = 10<br>N (%) | p-<br>value* |
| --- | --- | --- | --- | --- | --- |
| **Be required to participate in training when first employed** |  |  |  |  | - |
| Yes | 57 (78.1) | - | - | - |  |
| No | 16 (21.9) | - | - | - |  |
| **Non-required staff (n=16) believe training should be provided ‡** |  |  |  |  | - |
| Yes | 9 (56.3) | - | - | - |  |
| No | 7 (43.7) | - | - | - |  |
| **Required training frequency by employer** |  |  |  |  | - |
| Monthly | 9 (12.3) | - | - | - |  |
| Every 6 months | 6 (8.2) | - | - | - |  |
| Annually | 24 (32.9) | - | - | - |  |
| Don't know | 25 (34.2) | - | - | - |  |
| **Cultural safety** |  |  |  |  | .76 |
| No training received and no training needed | <5 (2.7) | <5 (5.3) | <5 (2.3) | 0 (0.0) |  |
| No training received but needed more training | 24 (32.9) | 5 (26.3) | 14 (31.8) | 5 (50.0) |  |
| Training received and needed more training | 23 (31.5) | 5 (26.3) | 15 (34.1) | <5 (30.0) |  |
| Training received and satisfied | 24 (32.9) | 8 (42.1) | 14 (31.8) | <5 (20.0) |  |
| **Aboriginal and/or Torres Strait Islander history** |  |  |  |  | .28 |
| No training received and no training needed | <5 (2.7) | <5 (5.3) | <5 (2.3) | 0 (0.0) |  |
| No training received but needed more training | 24 (32.9) | 5 (26.3) | 14 (31.8) | 5 (50.0) |  |
| Training received and needed more training | 26 (35.6) | 6 (31.6) | 15 (34.1) | 5 (50.0) |  |
| Training received and satisfied | 21 (28.8) | 7 (36.8) | 14 (31.8) | 0 (0.0) |  |
| **Aging and health needs** |  |  |  |  | .13 |
| No training received and no training needed | <5 (4.1) | 0 (0.0) | <5 (6.8) | 0 (0.0) |  |
| No training received but need training | 24 (32.9) | 8 (42.1) | 10 (22.7) | 6 (60.0) |  |
| Training received and needed more training | 27 (37.0) | 7 (36.8) | 16 (36.4) | <5 (40.0) |  |
| Training received and satisfied | 19 (26.0) | <5 (21.1) | 15 (34.1) | 0 (0.0) |  |
| **Social and emotional wellbeing** |  |  |  |  | .15 |
| No training received and no training needed | <5 (2.7) | 0 (0.0) | <5 (4.5) | 0 (0.0) |  |
| No training received but needed more training | 34 (46.6) | 9 (47.4) | 17 (38.6) | 8 (80.0) |  |
| Training received and needed more training | 15 (20.5) | 5 (26.3) | 8 (18.2) | <5 (20.0) |  |
| Training received and satisfied | 22 (30.1) | 5 (26.3) | 17 (38.6) | 0 (0.0) |  |
| **Healthy aging** |  |  |  |  | .07 |
| No training received and no training needed | <5 (4.1) | 0 (0.0) | <5 (6.8) | 0 (0.0) |  |
| No training received but needed more training | 30 (41.1) | 6 (31.6) | 16 (36.4) | 8 (80.0) |  |
| Training received and needed more training | 22 (30.1) | 9 (47.4) | 11 (25.0) | <5 (20.0) |  |
| Training received and satisfied | 18 (24.7) | <5 (21.1) | 14 (31.8) | 0 (0.0) |  |
| **Trauma-informed care** |  |  |  |  | .45 |
| No training received and no training needed | <5 (1.4) | 0 (0.0) | <5 (2.3) | 0 (0.0) |  |
| No training received but needed more training | 31 (42.5) | 7 (36.8) | 17 (38.6) | 7 (70.0) |  |
| Training received and needed more training | 29 (39.7) | 7 (36.8) | 19 (43.2) | <5 (30.0) |  |
| Training received and satisfied | 12 (16.4) | 5 (26.3) | 7 (15.9) | 0 (0.0) |  |
| **Grief and loss** |  |  |  |  | .42 |
| No training received and no training needed | <5 (2.7) | 0 (0.0) | <5 (4.5) | 0 (0.0) |  |
| No training received but needed training | 33 (45.2) | 8 (42.1) | 18 (40.9) | 7 (70.0) |  |
| Training received and needed more training | 20 (27.4) | 6 (31.6) | 11 (25.0) | <5 (30.0) |  |
| Training received and satisfied | 18 (24.7) | 5 (26.3) | 13 (29.5) | 0 (0.0) |  |
| **Dementia** |  |  |  |  | .58 |
| No training received and no training needed | <5 (2.7) | 0 (0.0) | <5 (2.3) | <5 (10.0) |  |
| No training received but needed training | 21 (28.8) | 7 (36.8) | 11 (25.0) | <5 (30.0) |  |
| Training received and needed more training | 28 (38.4) | 5 (26.3) | 19 (43.2) | <5 (40.0) |  |
| Training received and satisfied | 22 (30.1) | 7 (36.8) | 13 (29.5) | <5 (20.0) |  |
| **Aged care quality standards or indicators** |  |  |  |  | .002 |
| No training received and no training needed | 8 (11.0) | 0 (0.0) | <5 (6.8) | 5 (50.0) |  |
| No training received but needed training | 10 (13.7) | <5 (21.1) | <5 (9.1) | <5 (20.0) |  |
| Training received and needed more training | 30 (41.1) | 9 (47.4) | 18 (40.9) | <5 (30.0) |  |
| Training received and satisfied | 25 (34.2) | 6 (31.6) | 19 (43.2) | 0 (0.0) |  |

Data were presented as n (%) unless stated otherwise. \* Percentages were compared using Fisher's Exact test.
‡ 16 respondents who were not required to participate in training when first employed

Among respondents with no prior training, more than one-third expressed a need for training in social and emotional wellbeing (46.6%), grief and loss (45.2%), trauma-informed care (42.5%), and healthy ageing (41.1%). Additionally, over one-third of respondents who had previously received training reported a need for further training in Aged Care Quality Standards (41.1%), trauma-informed care (39.7%), dementia (38.4%), ageing and health needs (37.0%), and Aboriginal and Torres Strait Islander history (35.6%). Qualitative responses to ‘Other’ revealed training preferences in Aboriginal languages, mental health support, substance use, Advance Care Directives, and palliative care.

### 3.3 Training Attitude

Table 3 outlines training attitudes. Almost all respondents agreed or strongly agreed that training was important for skill development (98.6%), in delivering quality care (98.6%) and overall job satisfaction (83.6%). Additionally, 80.8% agreed or strongly agreed that they enjoy participating in training. However, fewer participants reported having received training specific to supporting Aboriginal and Torres Strait Islander peoples in aged care: just over half (52.1%) agreed or strongly agreed, while 39.7% disagreed or strongly disagreed.

**Table 3.**
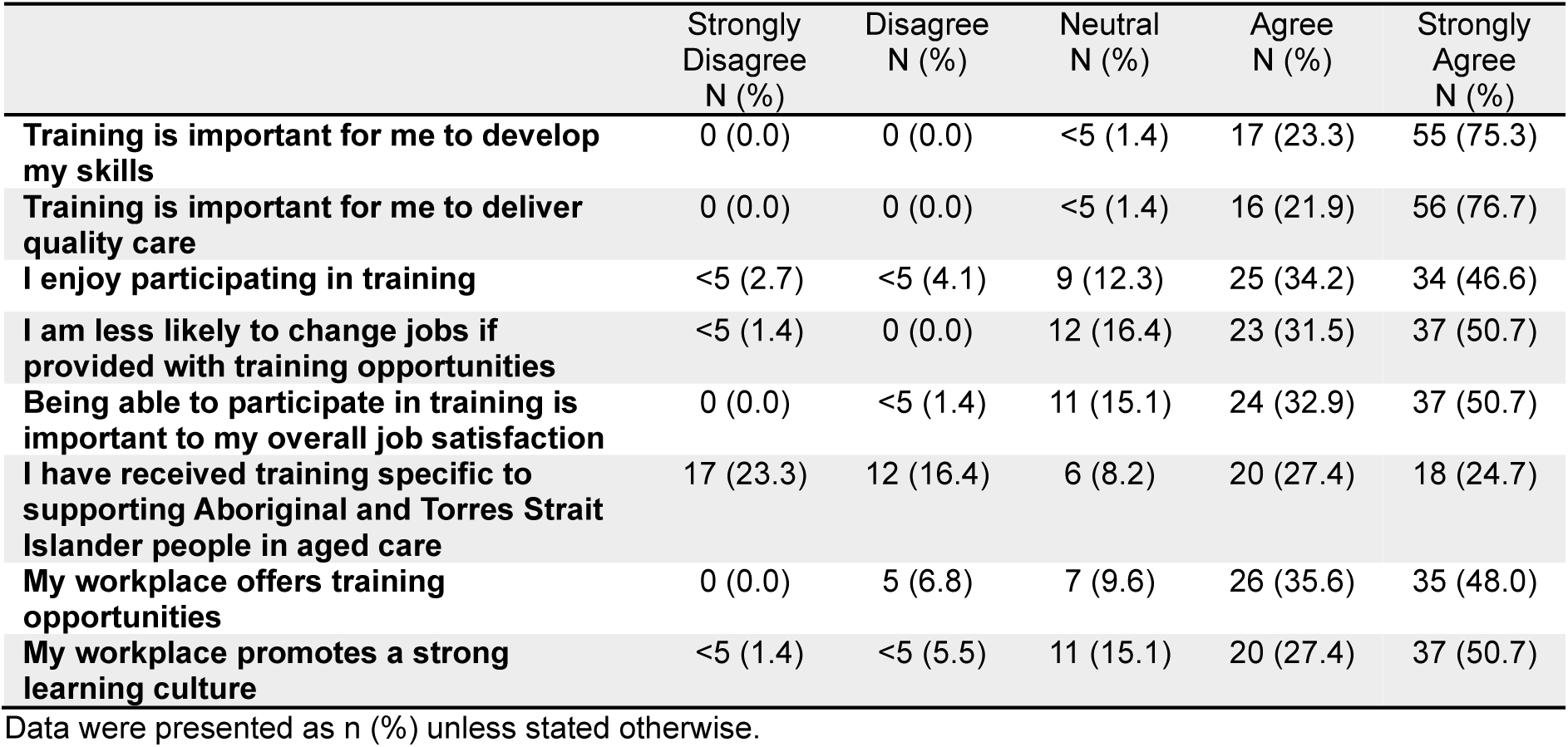
Training attitudes (n=73)

|  | Strongly Disagree<br>N (%) | Disagree<br>N (%) | Neutral<br>N (%) | Agree<br>N (%) | Strongly Agree<br>N (%) |
| --- | --- | --- | --- | --- | --- |
| **Training is important for me to develop my skills** | 0 (0.0) | 0 (0.0) | <5 (1.4) | 17 (23.3) | 55 (75.3) |
| **Training is important for me to deliver quality care** | 0 (0.0) | 0 (0.0) | <5 (1.4) | 16 (21.9) | 56 (76.7) |
| **I enjoy participating in training** | <5 (2.7) | <5 (4.1) | 9 (12.3) | 25 (34.2) | 34 (46.6) |
| **I am less likely to change jobs if provided with training opportunities** | <5 (1.4) | 0 (0.0) | 12 (16.4) | 23 (31.5) | 37 (50.7) |
| **Being able to participate in training is important to my overall job satisfaction** | 0 (0.0) | <5 (1.4) | 11 (15.1) | 24 (32.9) | 37 (50.7) |
| **I have received training specific to supporting Aboriginal and Torres Strait Islander people in aged care** | 17 (23.3) | 12 (16.4) | 6 (8.2) | 20 (27.4) | 18 (24.7) |
| **My workplace offers training opportunities** | 0 (0.0) | 5 (6.8) | 7 (9.6) | 26 (35.6) | 35 (48.0) |
| **My workplace promotes a strong learning culture** | <5 (1.4) | <5 (5.5) | 11 (15.1) | 20 (27.4) | 37 (50.7) |

Data were presented as n (%) unless stated otherwise.

High proportions of respondents indicated the importance of various training topics as “extremely important” (Table 4). Respondents with and without prior training identified key topics as cultural safety (80.8%), trauma-informed care (78.1%), social and emotional wellbeing (75.3%), grief and loss (74.0%), Aboriginal and Torres Strait Islander history (71.2%), ageing and health needs (67.1%), healthy aging (64.4%), and dementia (54.8%). A significant difference in attitudes towards training in aged care quality standards or indicators was observed by Indigenous status (p = .005, percentages compared using the Kruskal-Wallis test). Most Aboriginal respondents (61.4%) and most non-Aboriginal respondents (47.4%) rated this training topic as *extremely important*, whereas most respondents who preferred not to disclose their Indigenous status (60.0%) rated it as *not at all important*.

**Table 4.** Attitudes towards training topics (n=73)

|  | Not at all<br>important<br>N (%) | Slightly<br>important<br>N (%) | Neutral<br>N (%) | Very<br>important<br>N (%) | Extremely<br>important<br>N (%) |
| --- | --- | --- | --- | --- | --- |
| **Cultural safety** | 0 (0.0) | 0 (0.0) | 0 (0.0) | 14 (19.2) | 59 (80.8) |
| **Aboriginal and/or Torres Strait**<br>**Islander history** | 0 (0.0) | 0 (0.0) | <5 (1.4) | 20 (27.4) | 52 (71.2) |
| **Ageing and health needs** | 0 (0.0) | 0 (0.0) | <5 (2.7) | 22 (30.1) | 49 (67.1) |
| **Social and emotional wellbeing** | 0 (0.0) | 0 (0.0) | 0 (0.0) | 18 (24.7) | 55 (75.3) |
| **Healthy ageing** | 0 (0.0) | 0 (0.0) | 6 (8.2) | 20 (27.4) | 47 (64.4) |
| **Trauma-informed care** | 0 (0.0) | 0 (0.0) | <5 (1.4) | 15 (20.5) | 57 (78.1) |
| **Grief and loss** | 0 (0.0) | 0 (0.0) | <5 (4.1) | 16 (21.9) | 54 (74.0) |
| **Dementia** | 0 (0.0) | 0 (0.0) | <5 (5.5) | 29 (39.7) | 40 (54.8) |
| **Aged care quality standards or**<br>**indicators** | 12 (16.4) | 0 (0.0) | <5 (5.5) | 19 (26.0) | 38 (52.1) |

Data were presented as n (%) unless stated otherwise.

### 3.4 Barriers to Training and Preferences for Training Delivery Modes

Barriers to training and preferences for training delivery modes are presented in Table 5. *Time constraints* (58.9%) and *availability of training* (46.6%) were the two main barriers reported by respondents. 60.0% of respondents who preferred not to disclose their Indigenous status reported *other barriers*, compared with 26.3% of Aboriginal respondents and 13.6% of non-Aboriginal respondents (p = 0.01).

**Table 5.**
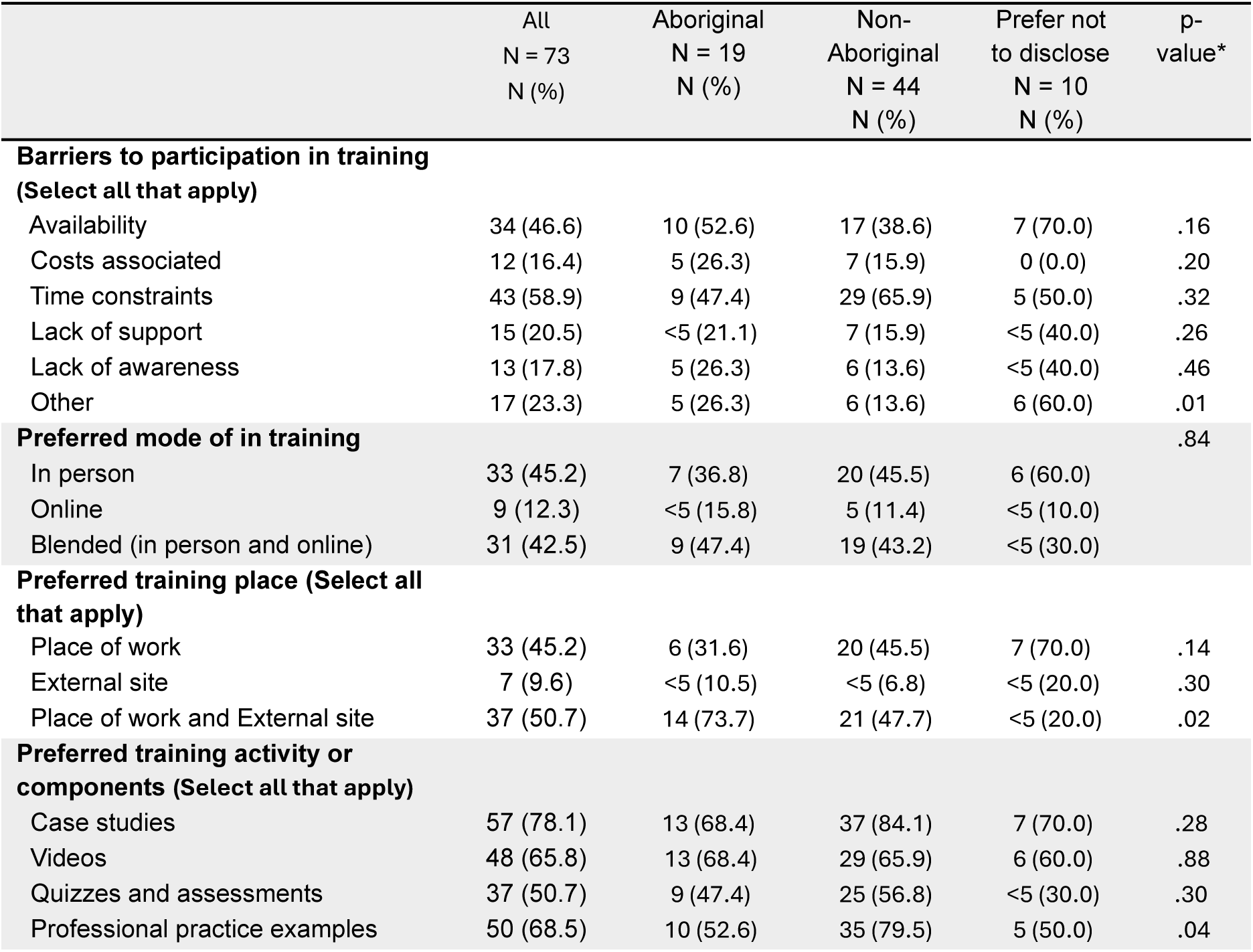
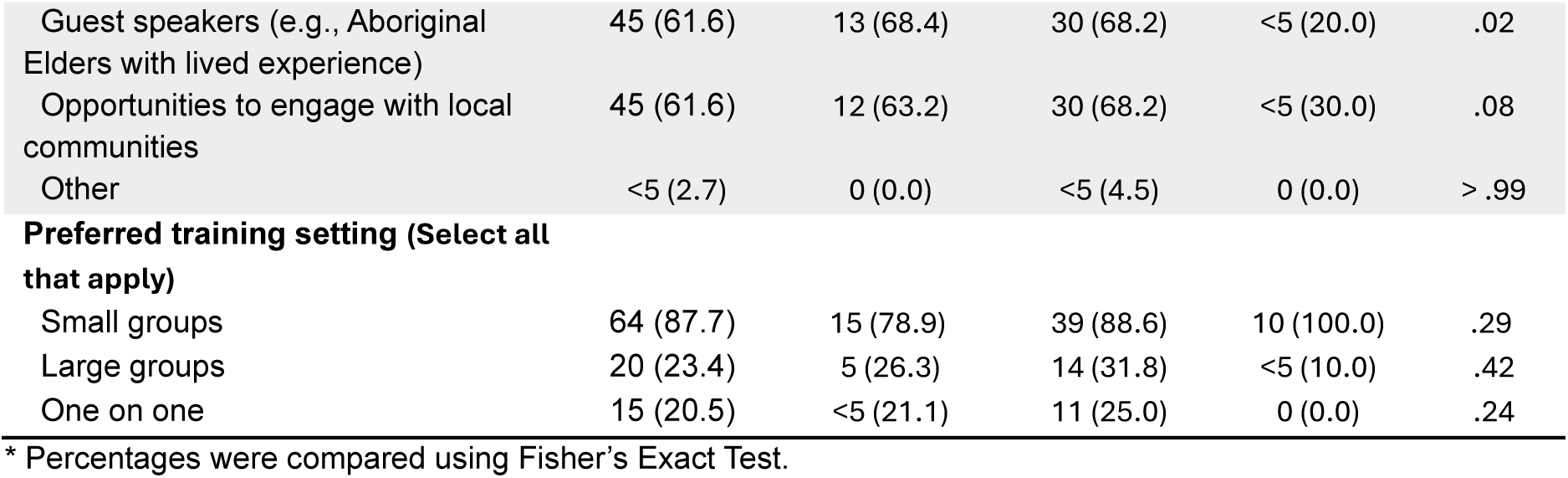
Barriers to training and preferences for training delivery modes.

*In-person training* was the most preferred delivery mode (45.2%), followed closely by *a blended approach of in-person and online* (42.5%), at their *place of work and external sites* (50.7%). Most Aboriginal respondents preferred to undertake training both in *workplace and external sites*, compared with 47.7% of non-Aboriginal respondents and only 20.0% of respondents who preferred not to disclose their Indigenous status (p = 0.02).

Among training components, the inclusion of *case studies* was the most favoured activity (78.1%) followed closely by *professional practice examples* (68.5%), *videos* (65.8%) and opportunities to *engage with the local community* and *guest speakers* (61.6% respectively). A significant difference was observed in preference for *professional practice examples* by Indigenous status (p = .04). 79.5% of non-Aboriginal respondent preferred this training activity, compared with 52.6% of Aboriginal respondents and 50.0% of respondents who preferred not to disclose their Indigenous status. Another significant difference was observed in preference for *guest speakers* by Indigenous status (p = .02). Most Aboriginal respondents (68.4%) and non-Aboriginal respondents (68.2%) preferred this training activity, compared with only 20.0% respondents who preferred not to disclose their Indigenous status. Most respondents also preferred to participate in *small group training* (87.7%).

## 4. Discussion

This study examined the training experiences, attitudes and needs of the aged care workforce providing services to Aboriginal and Torres Strait Islander peoples in South Australia. As the sector undergoes significant reform, including the introduction of the new Support at Home Program, strengthened Aged Care Quality Standards, and a new Aged Care Act,^23^ this workforce stands at the frontline of efforts to improve the experiences and aged care outcomes of Aboriginal and Torres Strait Islander peoples. The workforce influences not only the delivery of care, but also the extent to which Aboriginal and Torres Strait Islander peoples feel culturally safe, respected and recognised within the aged care system. Access to high-quality training that reflects the needs, aspirations and experiences of Aboriginal and Torres Strait Islander peoples is therefore essential. Without it, the aged care workforce cannot be expected to deliver the trauma-informed, relationship-based and culturally responsive care required by national reform agendas.^19^

While there were few significant differences in overall training needs, attitudes, and perceived importance of training, some meaningful differences emerged. We were able to identify some significant differences between Aboriginal and non-Aboriginal respondents and those who preferred not to identify their Indigenous status. This is important for ensuring training programs are appropriately tailored to the needs of Indigenous and non-Indigenous aged care workers, while also improving understanding of a sizeable segment of the workforce who may prefer not to disclose their ethnicity. Aboriginal respondents were more likely to work in community-controlled organisations, place higher importance on training about aged care quality standards and prefer delivery across both workplace and external sites. Preferences for training components also differed: non-Aboriginal workers favoured professional practice examples, whereas Aboriginal respondents valued guest speakers such as Elders, reflecting relational and culturally grounded learning approaches. These findings indicate that although shared core training needs exist across the aged care workforce, training must remain flexible and culturally responsive to the distinct contexts and responsibilities of Aboriginal workers, particularly those in community-controlled and remote environments.

Notably, the training areas of greatest unmet need identified in this study – social and emotional wellbeing, trauma-informed care, grief and loss, and healthy ageing – were those identified by Aboriginal and Torres Strait Islander peoples as essential knowledge areas for the aged care workforce within previous research.^14–16,18^ The need for training in these topics (with most rated as “extremely important”) demonstrates not only unmet training need but is consistent with a shared workforce recognition of the importance of holistic, culturally grounded approaches to Aboriginal and Torres Strait Islander ageing and wellbeing.^17,24^ However, this sits in stark contrast to current training provision: fewer than half of respondents had received any training specific to supporting Aboriginal and Torres Strait Islander peoples in aged care. Taken together, these findings underscore a critical disconnect between what communities consider important for high-quality, culturally safe care and the scope of training currently available to the workforce.

Importantly, these findings should not be interpreted as reflecting deficiencies within individual organisations providing aged care services. Across the sector, workforce training capacity is shaped by broader structural and system-level constraints, including funding models that do not adequately resource training, workforce shortages, high service demand, and increasing regulatory and reporting requirements.^15,16^ This is particularly evident within the Aboriginal community-controlled sector, which often operate under compounding resourcing constraints.^25,26^ Responsibility for ensuring a capable, culturally safe aged care workforce therefore extends beyond individual providers and sits across governments, regulators, training bodies, and commissioning frameworks.

Our findings also provide insight into how the aged care staff prefer to learn. Respondents overwhelmingly favoured small-group formats, case studies and real-world practice examples, alongside opportunities to engage with community and with Aboriginal guest speakers. In-person or blended delivery was strongly preferred, underscoring the value placed on relational, reflective and experiential learning.^27–29^ At the same time, the results further illustrate persistent barriers that continue to limit workforce participation in training. Consistent with the broader literature,^15,27,30,31^ respondents in this study identified time constraints, limited availability, and inadequate organisational support as key barriers to participating in training. These challenges may be intensified by workforce shortages^3^ and growing service demand^4^ which may further reduce opportunities for the workforce to upskill. Embedding training as a core and resourced component of workforce planning, rather than an optional or compliance driven activity, is therefore essential.

Collectively, these findings reinforce the urgent need for a coordinated and culturally led approach to workforce development within aged care. While the Royal Commission into Aged Care Quality and Safety^7^ and the Interim First Nations Aged Care Commissioner’s report^10^ have both highlighted the systemic failures that leave Aboriginal and Torres Strait Islander peoples feeling unsafe and disempowered within aged care, this study extends these insights by identifying how reform can be operationalised through increased workforce training. Our findings extend this evidence base by demonstrating that the gaps identified at the system level are mirrored at the workforce level, where training remains sporadic, poorly targeted, and insufficiently aligned with the realities of Aboriginal and Torres Strait Islander ageing.

Future aged care training programs must therefore be responsive to the diverse roles and settings across the sector and designed with flexibility, accessibility, and cultural integrity in mind. Expanding beyond their current remit, training programs must equip workers with the knowledge, skills and confidence to deliver care that is trauma-informed, culturally safe and responsive to the holistic needs of Aboriginal and Torres Strait Islander peoples.^1,15^ Partnerships with Aboriginal community-controlled organisations are central to achieving this vision. Such partnerships ensure that training reflects local contexts, draws on community authority, and positions Aboriginal and Torres Strait Islander peoples as experts and leaders in curriculum design, delivery, and evaluation. Building curricula that foreground Aboriginal and Torres Strait Islander knowledges, experiences, and needs, and that critically engage with the social and cultural determinants of health, is essential for achieving equity, dignity, and trust in aged care.^15–18^

The findings of this study have directly informed the development of *Walking Together in Aged Care,* a training program dedicated to building the capacity of the aged care workforce in meeting the needs of Aboriginal and Torres Strait Islander peoples. In particular, the findings guided training topic selection and content, in addition to delivery modes and interactive activities. By clarifying where capability gaps exist, how staff experience current training, and what delivery approaches best support learning, this study offers practical guidance for strengthening workforce development across the sector. Addressing the identified gaps and barriers will be essential to building a culturally safe, confident and well-supported aged care workforce capable of meeting the needs of older Aboriginal and Torres Strait Islander peoples.

## 5. Strengths and limitations

This study is, to our knowledge, the first to explore the perspectives of the aged care workforce involved in the direct care (e.g., personal care, allied health) for older Aboriginal and Torres Strait Islander peoples.^15^ It was developed in response to the identified needs of our partner organisation, ACS, who are the largest provider of aged care services for Aboriginal and Torres Strait Islander peoples in South Australia, ensuring strong relevance and practical application. Additionally, the study sample comprised only those working with Aboriginal and Torres Strait Islander clients, providing a clear understanding of workforce training needs in these contexts. However, the study’s findings should be interpreted considering several limitations. As total workforce numbers of each organisation invited to do the survey were not collected, the representativeness of the sample cannot be determined, and findings may not be generalisable to the broader aged care workforce. The study was also limited to South Australia and reflects the workforce context, service structures and demographic profile of this jurisdiction; therefore, findings may not capture variation across other states and territories or the experiences of aged care workers operating in different policy, cultural or organisational environments. We did not recruit from state government providers who could be major providers in regional areas, due to the short time frame of the study and the additional ethical approvals required to access these services. Additionally, the cross-sectional design did not allow for the assessment of content, quality, or effectiveness of the training received by respondents.

## 6. Conclusion

This study provides critical insight into the training experiences, needs and preferences of the aged care workforce supporting Aboriginal and Torres Strait Islander peoples. The findings reveal a motivated workforce with strong commitment to culturally safe care, yet one that remains constrained by limited access to culturally specific training, inconsistent and/or unclear organisational expectations, and structural barriers that restrict participation. Priority training areas identified align closely with what Aboriginal and Torres Strait Islander communities have long identified as essential for high-quality care. Preferences for small-group, experiential and community-led learning further highlight the need for training models grounded in relational, contextually relevant pedagogy. Despite the *Royal Commission into Aged Care Quality and Safety* calling for improvements in the provision of workforce training within Aboriginal and Torres Strait Islander aged care, our results demonstrate a continued lack of training specific to supporting Aboriginal and Torres Strait Islander people in aged care. Fully realising the potential benefits of dedicated professional training programs across the aged care sector requires the necessary policy and fiscal investments.

## Data Availability

Data from this study are not available due to the sensitive nature of the content discussed.

## Acknowledgements

The authors would like to acknowledge and pay their respects to the Traditional Owners of the lands on which this research was conducted. We pay our respects to Elders past, present and emerging. We extend our gratitude to study participants’ for sharing their experiences and views, our partnership with ACS, and the PSC who provided governance and advice.

## Availability of data and materials

Data from this study are not available due to the sensitive nature of the content discussed.

